# Sex-specific Axonal Conduction Velocity Development Drives Differential Changes in Frontal, Parietal, and Insular Cortices in Autism Spectrum Disorder

**DOI:** 10.1101/2025.10.28.25338995

**Authors:** Benjamin T. Newman, Haylee J. Ressa, Zachary Jacokes, James C. McPartland, Natalia M. Kleinhans, Sara Jane Webb, Abha R. Gupta, Raphael A. Bernier, Nadine Gaab, Allison Jack, T. Jason Druzgal, Kevin A. Pelphrey, John Darrell Van Horn, the GENDAAR Research Consortium

**Author notes:** These authors are co-first authors of this manuscript (Benjamin T. Newman, Haylee J. Ressa, John Darrell Van Horn).

## Abstract

Neither the neurological underpinnings of autism spectrum disorder (ASD) nor their contribution to sex differences are well understood. In previous cross-sectional studies of axonal conduction velocity, the speed of action potential transmission, was observed to be decreased in autistic individuals, and this deficiency was associated with cognitive and behavioral differences. This longitudinal study aims to better understand how changes in neuronal microstructure contribute to the developmental trajectory of individuals with ASD and specifically to sex-differences in behavior during the adolescent period. Eighty-two participants (34 ASD, 41 female) completed multi-year longitudinal behavioral and neuroimaging testing. Pubertal development significantly mediated and accelerated age-related increases in conduction velocity, with girls with autism exhibiting greater increases in cortex over time and boys exhibiting greater increases in white matter (WM). Girls with autism exhibited more rapid increases in frontal and parietal cortices while boys showed relatively higher increases in insular cortex compared to girls. Across all boys, conduction velocity increased in WM at a higher rate than girls, but increased more slowly in autistic relative to non-autistic boys. Parent-reported anxious and depressive symptomatology also increased over time in girls with autism, whereas behavioral metrics associated with ASD declined, especially in boys. Notably, conduction velocity showed significant associations with parent-reported anxious and depressive symptomatology in many of the same brain regions that showed sex-specific developmental changes. These results indicate that neurodevelopmental changes in conduction velocity may underlie sex-linked biological mechanisms and contribute to differences in behavioral expression in autistic and non-autistic development.

## Introduction

Autism spectrum disorder (ASD) is characterized by differences in behavior, including challenges in social communication and a tendency toward restrictive and repetitive behaviors and interest (Masi et al., 2017; Regier et al., 2013). ASD encompasses a highly diverse spectrum of behavioral profiles and severity, with some individuals requiring minimal support and others requiring lifelong services along different language, social, and behavioral dimensions (Lord et al., 2020). These behavioral profiles can change as children with ASD develop into adults, especially with targeted and sustained therapies and intervention. Additionally, significant differences exist in trait patterns by sex (Braden et al., 2021). In particular, girls with ASD may present with fewer externalizing behaviors, such as aggression and hyperactivity, than boys, a relative difference that has been theorized to contribute to the underdiagnosis of girls (White et al., 2017). Understanding the heterogeneity of trait expression is crucial to predicting the trait patterns of an individual across their lifetime and providing optimal support and individually tailored interventions.

A diagnosis of ASD typically involves a comprehensive evaluation using reliable standardized measurers of behavior performed by a clinician, such as the Autism Diagnosis Observation Schedule (ADOS) and a clinical interview covering early developmental (Lord et al., 1994, 2000). Additional behavioral tests used to describe adolescent behavior and well-being, while not diagnostic in nature, frequently display differences between ASD and non-ASD populations. However, the connection between these behavioral test metrics, especially specific components or subscales of each test that contribute to the diagnosis, and the neurological basis of ASD remains uncertain. Finding neural correlates across development that align with behavioral variability has the potential to inform predictive biomarkers, which in turn may inform clinical practice, particularly on co-occuring features such as anxiety or depression. This study specifically aims to characterize how longitudinal changes in cellular microstructure relate to behavioral variability in adolescents with ASD, especially how cellular microstructure differences may relate to sex-specific behavioral differences. Identifying sex-specific developmental patterns in white matter structure can advance development of biomarkers that capture the heterogeneity of ASD across the lifespan.

Atypical white matter microstructure has been associated with the neurobiology of ASD in many previous studies, particularly in regions such as the corpus callosum (Travers et al., 2012; Zhao et al., 2022) and frontal cortex (Dimond et al., 2019). White matter microstructure characteristics, like axonal diameter, density, and myelination, can be studied using diffusion magnetic resonance imaging (dMRI), a neuroimaging technique sensitive to water molecules’ movement (Afzali et al., 2021). As deficiencies in axonal structure, such as irregular diameter or disrupted myelination, may impair the speed or efficiency of action potentials, axonal characteristics can be important factors in signal transduction (Liewald et al., 2014). In multiple major white matter tracts, adolescents and young adults with ASD have been found to have reduced fiber density, a measure of the number of nerve fibers, or axons, present in an area(Dimond et al., 2019). Furthermore, lower fiber density, specifically in the splenium of the corpus callosum, was found to be significantly associated with increased social impairment in participants with ASD (Dimond et al., 2019). In adults, however, lower fiber density in the corpus callosum was only found in female participants with ASD and not male participants (Kirkovski et al., 2022). This pattern may suggest that fiber density normalizes by adulthood in male but not female autistic individuals (Braden et al., 2021). Investigating these microstructural differences in longitudinal cohorts is an essential next step in advancing these findings.

This investigation is particularly crucial as puberty is a period of rapid microstructural change both in terms of axonal refinement and increased myelination across major tracts (Lebel & Deoni, 2018). Longitudinal diffusion imaging has shown that puberty may drive sex-specific increases in white matter integrity, demonstrating its potential role in shaping cognitive and behavioral trajectories during adolescence (Simmonds et al., 2014). Puberty further has been shown to act on glial cells, increasing myelination throughout the WM and innervating the cortex (Piekarski et al., 2023). At a microstructural level it has previously been shown that this myelinating process increases intracellular signal throughout the WM skeleton, suggesting that glial reactivity to pubertal hormones affects axonal environments throughout the brain and across adolescence (Newman et al., 2023).

Architectural differences in the axon microstructure have additionally been observed in participants with ASD in recent work focusing on measurements of conduction velocity and g-ratio (Newman et al., 2024). Conduction velocity quantifies the speed at which an action potential travels along an axon, reflecting the axon’s capacity to transmit information and is sensitive to variations in myelination and axonal development. Conduction velocity is derived in part from g-ratio, a proportion of axonal diameter to myelin diameter with an optimal range of around 0.6-0.7 (Rushton, 1951). Differences in conduction velocity, g-ratio, and extracellular water were found in ASD participants in early adolescence and were present throughout the cortex, subcortex, and white matter skeleton. In particular, reduced g-ratio and conduction velocity in ASD participants may indicate potential impairments in long-range connections, which depend on thicker axons and myelin for efficient signal transmission (Newman et al., 2024). Further research in our group has demonstrated that g-ratio and conduction velocity are significantly associated with a wide variety of social, behavioral, and cognitive measures in ASD and suggest that age-related maturation of brain metrics may drive changes in ASD behavior. If ASD can be categorized into subtypes, these distinct groups may exhibit varying behavioral trajectories throughout development (Ressa et al., 2024).

To continue this work, this study leverages a rich longitudinal dataset including a cohort of autistic and non-autistic individuals from the NIH-sponsored (MH100028) Autism Centers of Excellence (ACE). This cohort consists of a large and carefully evaluated group of age- and sex -matched individuals in which we can explore the relationship between brain microstructure and behavioral patterns. We investigated age-related maturation of brain metrics and differences that arise throughout development between ASD and neurotypical individuals. We also examined sex differences in these developmental patterns. In addition, we compared behavioral metrics across two timepoints within the ASD cohort, stratified by sex. This approach allowed us to identify which behavioral measures are most indicative of sex differences over time and to refine the scope of subsequent microstructural analysis. Using the results of these broad analyses, we then focused on key findings that were significant both in terms of associations between behavior and brain metrics and in regard to sex-based differences in expression.

This study represents one of the first longitudinal investigations of brain microstructure in the context of ASD to further understanding of microstructure changes throughout adolescence. Ultimately, these efforts will advance our knowledge of how ASD progresses across development and support development of predictive biomarkers and personalized therapeutic supports.

## Methods

### Participants

Participant demographics are summarized in Table 1. 199 participants completed behavioral and cognitive testing at the baseline study visit. The average age of this sample was 12.85 years ± 2.93 S.D. at baseline and 18.17 years ± 2.95 S.D. at follow-up, and 49% were female. The ASD group consisted of 100 participants with a mean age of 12.74 ± 2.85 S.D. years at baseline of whom 47% were female. The control group included 99 participants with a mean age of 13.25 ± 2.97 S.D. years at baseline and 50.5% were female. Of the 199 participants, 82 returned for the follow-up visit and completed both behavioral and cognitive assessments as well as neuroimaging to be included in the final cohort. The mean age of this longitudinal cohort was 18.17 ± 2.88 S.D. years at follow-up and 50% were female. This cohort consisted of 34 individuals with ASD and 48 neurotypical controls. Three participants (2 ASD, 1 non-ASD, all male) were excluded from neuroimaging analysis due to scanning errors. To confirm demographic similarity between cohorts, we performed chi-squared tests for sex and diagnoses (ASD vs. control) and a t-test for age. There were no significant differences between groups on any of these variables (all p > 0.2), indicating that the samples were demographically comparable.

**Table 1:**
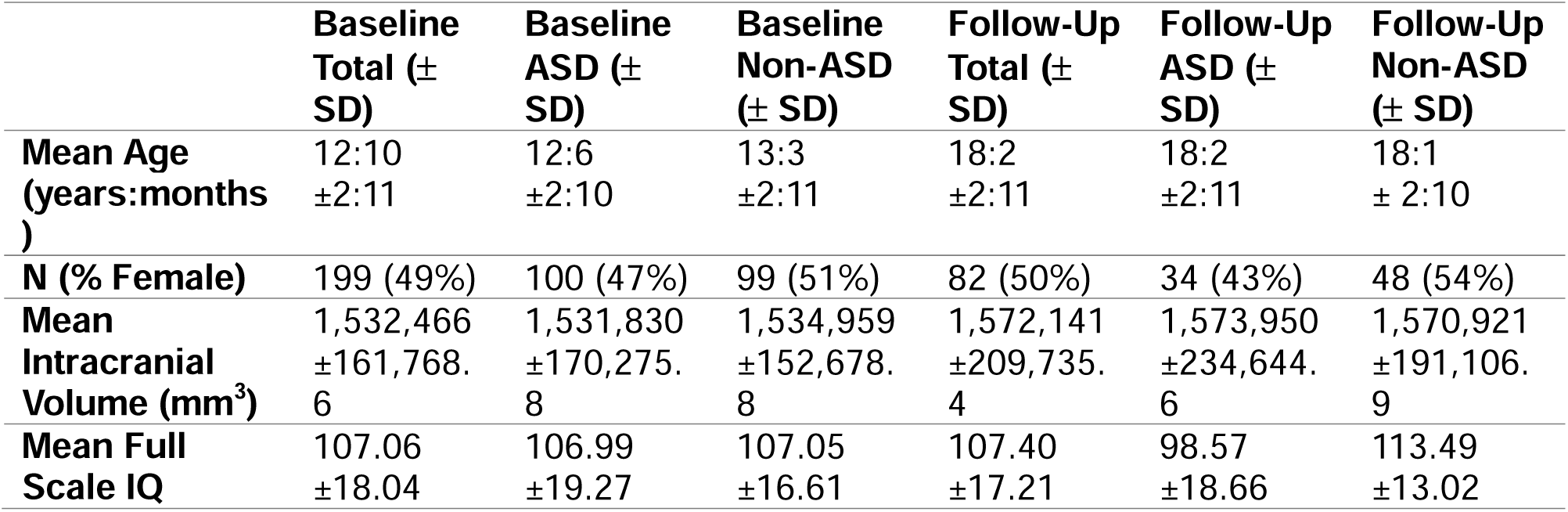
Demographics of study participants separated by diagnosis and timepoint.

|  | **Baseline**<br>**Total (±**<br>**SD)** | **Baseline**<br>**ASD (±**<br>**SD)** | **Baseline**<br>**Non-ASD**<br>**(± SD)** | **Follow-Up**<br>**Total (±**<br>**SD)** | **Follow-Up**<br>**ASD (±**<br>**SD)** | **Follow-Up**<br>**Non-ASD**<br>**(± SD)** |
| --- | --- | --- | --- | --- | --- | --- |
| **Mean Age**<br>**(years:months**<br>**)** | 12:10<br>$\pm 2:11$ | 12:6<br>$\pm 2:10$ | 13:3<br>$\pm 2:11$ | 18:2<br>$\pm 2:11$ | 18:2<br>$\pm 2:11$ | 18:1<br>$\pm 2:10$ |
| **N (% Female)** | 199 (49%) | 100 (47%) | 99 (51%) | 82 (50%) | 34 (43%) | 48 (54%) |
| **Mean**<br>**Intracranial**<br>**Volume (mm<sup>3</sup>)** | 1,532,466<br>$\pm 161,768$.<br>6 | 1,531,830<br>$\pm 170,275$.<br>8 | 1,534,959<br>$\pm 152,678$.<br>8 | 1,572,141<br>$\pm 209,735$.<br>4 | 1,573,950<br>$\pm 234,644$.<br>6 | 1,570,921<br>$\pm 191,106$.<br>9 |
| **Mean Full**<br>**Scale IQ** | 107.06<br>$\pm 18.04$ | 106.99<br>$\pm 19.27$ | 107.05<br>$\pm 16.61$ | 107.40<br>$\pm 17.21$ | 98.57<br>$\pm 18.66$ | 113.49<br>$\pm 13.02$ |

### Ethical Approval

Data from the ACE GENDAAR project used in this study were collected across four sites: Yale University, Harvard University/Boston Children’s Hospital, the University of California, Los Angeles, and the University of Washington/Seattle Children’s Research Institute. All procedures were approved by the institutional review boards of participating sites and conducted in accordance with the Declaration of Helsinki. The study received institutional review board (IRB) approvals as follows: Yale as lead institution (IRB: 1206010363; Pelphrey), UCLA (IRB# 10-000387-CR-00008; Bookheimer), Boston Children’s Hospital (IRB-P00004852; Nelson), and with Seattle Children’s Hospital (IRB 00000277; Webb) as a reliant site to Yale. George Washington University (GWU) assumed the role of lead institution (IRB 031802; Pelphrey) and the ACE project is now governed as a single-site IRB at the University of Virginia (UVA; IRB-HSR-220423; Pelphrey). The ACE GENDAAR Data Coordination Center (DCC) was originally governed by UCLA, subsequently by the University of Southern California (USC; HS-18-00467, HS-13-00668; Van Horn), and presently by UVA (IRB-HSR-22078; Van Horn). All de-linked phenotypic, neuroimaging, EEG, and genetics data from ACE GENDAAR has been shared with the NIMH Data Archive (NDA), beginning 11/23/2012, under study collection ID 2021 where they are available for download and re-analysis by NDA-approved investigators.

### MRI Image Acquisition

T1-weighted and diffusion images were acquired for all the participants. Diffusion images were acquired with an isotropic voxel size of 2×2×2mm^3^, 64 non-colinear gradient directions at b = 1000 s/mm2, and 1 b = 0, TR = 7300ms, TE = 74ms. T1-weighted MPRAGE images with a FOV of 176×256×256 and an isotropic voxel size of 1×1×1mm3, TE = 3.3; T2-weighted images were acquired with a FOV of 128×128×34 with a voxel size of 1.5×1.5×4mm3, TE = 35ms.

### Aggregate G-Ratio and Conduction Velocity Calculation

G-ratio is the ratio of axon diameter (d) to fiber diameter (D), as demonstrated by Equation 1. The fiber density cross section was used as the intra-axonal volume (AVF), and the T1w/T2w ratio was used as the myelin volume fraction (MVF). This equation was used on a voxel-wise basis according to the principles defined by Stikov et al. and Mohammadi & Callaghan (Mohammadi & Callaghan, 2021; Stikov et al., 2015).

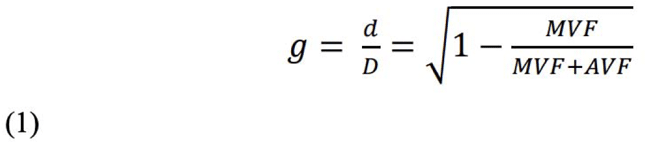

Aggregate conduction velocity was calculated according to Rushton and Berman (Berman et al., 2019; Rushton, 1951). Conduction velocity (θ) depends on the length of each fiber segment (l), which scales with the outer diameter of the fiber (D). The outer diameter (D) can be related to the axon diameter (d) and the g-ratio (g) according to Equation 2.

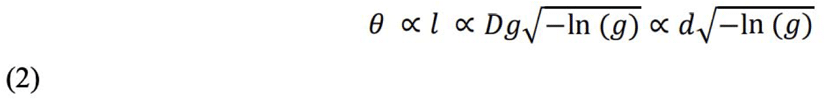

The mean value of these metrics was measured within each of the 164 regions of the Destrieux Cortical Atlas and 48 regions of the Johns Hopkins University White Matter Regions of Interest Atlas (Destrieux et al., 2010; Mori et al., 2005). There was additionally 2 summary ROIs made from each atlas, with a whole cortex ROI being created from all cortical ROIs in the Destrieux Cortical Atlas and a whole WM ROI being created from all 48 ROIs in the JHU-WM Atlas.

### Motion Assessment

Head motion during dMRI acquisition was quantified for each participant as average framewise displacement (in mm) prior to motion correction performed as part of FSL’s *eddy*. To assess whether motion differed by diagnostic group or sex, linear regression models were used to evaluate the relationship between motion and conduction velocity (Fig. 1). Across all cortical ROIs, motion was not significantly associated with mean cortical conduction velocity (β = –0.00247, p = 0.112), nor did group membership significantly predict conduction velocity (p = 0.208). Similar results were observed in the composite white matter ROI (β = –0.1739, p = 0.123). No significant differences in motion were observed between male and female participants (β = 0.1467, p = 0.186). These findings suggest that head motion is unlikely to confound observed group differences in conduction velocity or related metrics and was thus not included in any further analysis.

**Figure 1:**
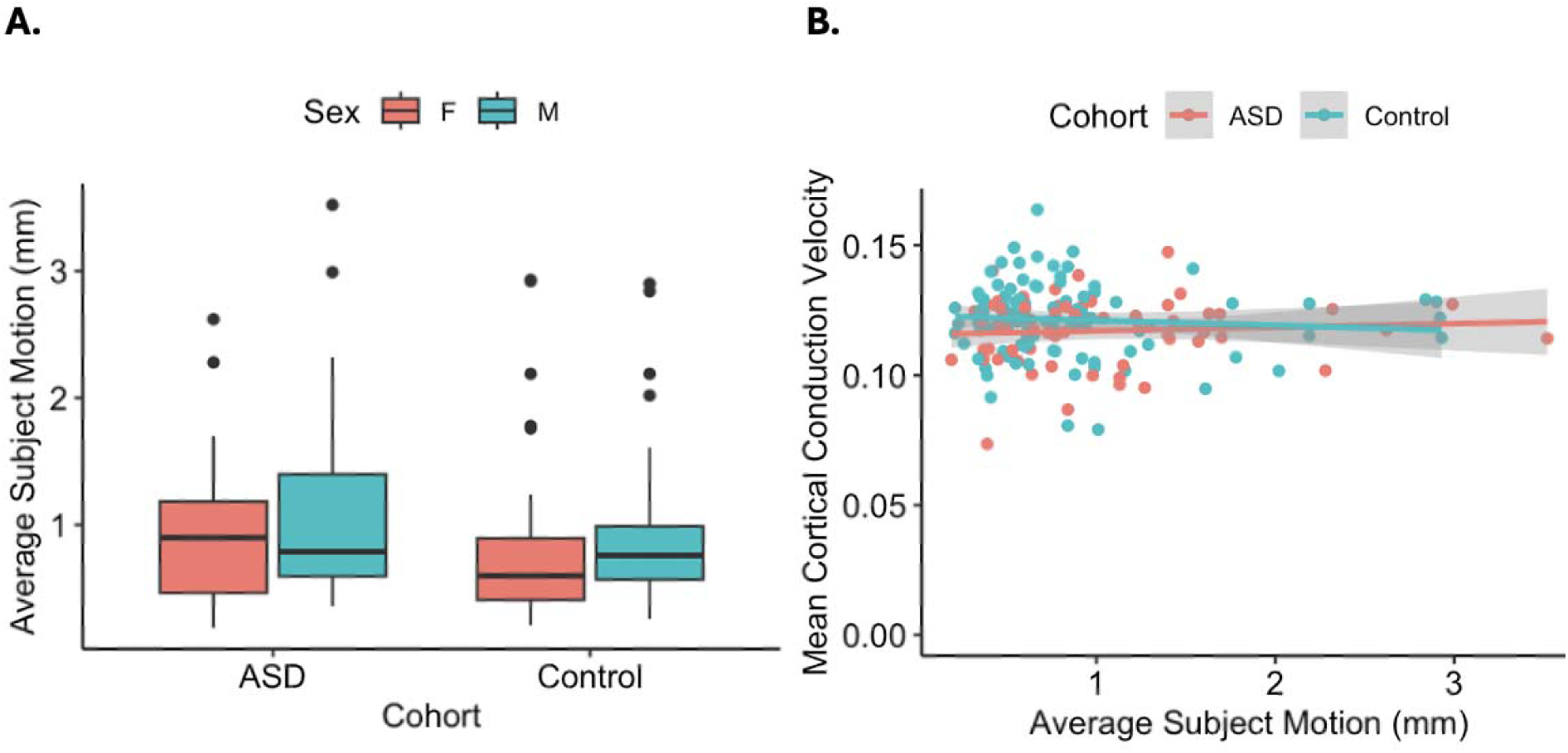
Motion and cortical conduction velocity. (A) Average head motion by sex and diagnostic group, showing no significant differences. (B) Scatterplot of mean cortical conduction velocity versus motion, with no significant associations observed in either group.

### Behavioral and Cognitive Assessments

ACE GENDAAR participants included male and female youth aged 8–17 years assigned to either the ASD or typically developing (TD) group. Written informed consent was obtained from parents and assent from children. General exclusion criteria included full-scale IQ (FSIQ) ≤ 70 (as estimated by the Differential Ability Scales–Second Edition General Conceptual Ability score), twin status, active tic disorder interfering with imaging, pregnancy, presence of metal in the body, seizures within the past year, or current use of benzodiazepines, barbiturates, or anti-epileptic medications. Use of other medications was permitted only if dosage and regimen were stable for ≥6 weeks.

### Typically Developing (TD)

Additional exclusionary criteria specific to the TD group were as follows: diagnosed, referred, or suspected ASD, schizophrenia, intellectual disability, learning disability, or other developmental or psychiatric disorder; a first-or second-degree relative with ASD; a total t-score > 60 on the Social Responsiveness Scale, Second Edition (Constantino & Gruber, 2012); a raw score > 11 on the Lifetime version of the Social Communication Questionnaire (Rutter, 2003); clinical impression suggesting ASD, other developmental delay or disorder, broader autism phenotype, or significant psychiatric disorder.

### Autism Spectrum Disorder (ASD)

Additional exclusionary criteria specific to the ASD group were as follows: known single gene disorder related to ASD or syndromic form of ASD (e.g., Fragile X); medical conditions likely to be etiological (e.g. focal epilepsy or infantile spasms); any neurological disorder involving pathology above the brainstem, other than uncomplicated non-focal epilepsy; history of significant pre-or perinatal injury, i.e. birth at < 36 weeks and weight < 2000 grams, or neonatal intensive care unit hospital stay > 3 days; history of neonatal brain damage; any known environmental circumstances that might account for the picture of ASD in the proband (e.g., severe nutritional or psychological deprivation); clinically significant visual or auditory impairment after correction; or any sensorimotor difficulties that would preclude valid use of the diagnostic instruments.

Children and adolescents who received the Autism Diagnostic Observation Schedule, Second Edition (ADOS-2) Module 3 (for which revised algorithms were available from the outset of the study) were required to achieve a Calibrated Severity Score (CSS) of _≥_ 4; adolescents who received Module 4 (for which a revised algorithm became available during the course of data collection (Hus & Lord, 2014)) were required to meet ASD criteria according to either the updated algorithm (CSS _≥_ 4) or the version of the algorithm published with the ADOS-2 (Communication + Social Interaction Total _≥_ 7). Scores on the Autism Diagnostic Interview-Revised (ADI-R), a standardized parent interview designed to obtain ASD symptom information, were required to meet the diagnostic algorithm within one point.

During both study waves, participants were administered an array of behavioral and cognitive tests listed in Supplementary Table 1. The following behavioral tests were used:

The Behavior Rating Inventory of Executive Functions (BRIEF) is a behavior rating scale used to screen for executive function deficits in children ages 5 to 18 years. Parents and teachers complete the BRIEF questionnaire, which asks how the child behaves in everyday situations, particularly those that require problem-solving. Children with ASD have been found to have significantly elevated BRIEF scores compared to neurotypical children, corresponding to executive function ability deficits (Blijd-Hoogewys et al., 2014).

The Child Behavior Checklist (CBCL) component of the Achenbach System of Empirically Based Assessment assesses a range of behavioral and emotional syndromes, including anxiety, depression, aggression, and defiant behavior problems. The syndromes are grouped into internally and externally focused behaviors and emotions. The CBCL questionnaire is administered to parents of children ages 6 to 18 years. Children with ASD have been found to have higher scores on CBCL subscales for depression, social problems, thought problems, and attention problems compared to neurotypical children (Arias et al., 2022). The Adult Behavior Checklist (ABCL) is a version of the CBCL that asks age-appropriate questions to assess participants over 18 years of age. The ABCL subscales are comparable to those of CBCL and were used for the adult participants in the second wave of the study.

The Social Responsiveness Scale Second Edition (SRS-2) is a rating scale measuring behaviors associated with ASD. It can be completed by raters with at least one month of experience with the rated individual. Different rating forms are available for age groups, including a self-report form for individuals aged 19 years and up. The SRS-2 focuses on social differences, and each item is responded to with a 4-point Likert scale rating (Bruni, 2014; Constantino & Gruber, 2012).

The Vineland Adaptive Behavior Scales – II (Vineland-II) is utilized to assess adaptive functioning across various domains. In contrast to other behavioral components measured in this study, Vineland-II aims to understand typical behavior in ‘real life’ situations outside the laboratory setting (Doll, 1935).

The Pubertal Development Scale (PDS) is a parent-reported scale that quantifies a participant’s estimated puberty development (Morningstar & Burns, 2025). The PDS questions are sex-specific and assess physical markers of puberty (such as voice deepening in males and onset of menses in females) on a scale from 1 to 4, with 4 being the most developed. The PDS was used to control for the earlier age of puberty onset typically seen in girls compared to boys.

### Statistical Approach

This study utilized a sequential 3 experiment approach with each analysis informing the subsequent model. For experiment 1, all behavioral, cognitive, and longitudinal test results were compared to the mean microstructural values in each ROI using general linear models. All resulting p-values were corrected for multiple comparisons using the Benjamini & Hochberg method (Benjamini & Hochberg, 1995) across all 214 ROIs. After FWE correction, no ROI was significant for two-or three-way interactions among diagnosis, sex, and time-point, so these interactions were discarded. After examining longitudinal change between timepoints, the following statistical model was used in experiment 1, with both the base longitudinal model and subsequent behavioral metric model including a non-interaction term for participant sex plus control terms (participant age, intracortical volume (ICV), scanner site, timepoint, and IQ) selected for scientific merit informed by previous microstructural analyses:

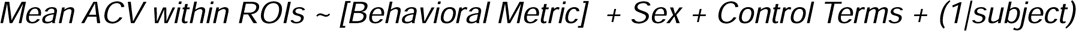

Due to finding few significant relationships between ACV and behavioral metrics longitudinally, in experiment 2 a mediation analysis was used to test whether pubertal development (PDS) mediated the relationship between age and conduction velocity. While in experiment 3 two models were used, a first model to select behavioral metrics with significant sex interaction across the longitudinal timeframe, including PDS following the mediation analysis:

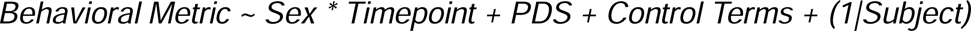

This was followed by an analysis using a second model to specifically test if the single significant behavioral metric was significantly related to ACV across the 214 ROIs:

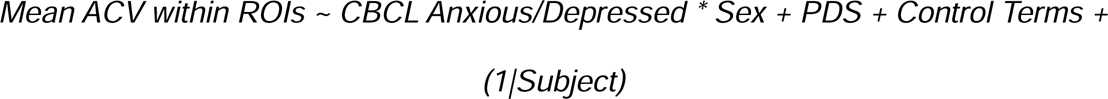

## Results

### Experiment 1

Age-related patterns of change in conduction velocity across both cortex and WM are visualized in Figure 2. The mean differences between baseline and follow-up of aggregate conduction velocity measured within all the cortical Destrieux ROIs and all Johns Hopkins University White Matter Regions of Interest are displayed in Figures 3 & 4. In the cortex in male participants, conduction velocity was significantly reduced in ASD compared to non-ASD males (T=2.476, p<0.05), but there was not a significant change in conduction velocity overall (T=1.733, p=0.088 n.s.) likely driven by the lack of change in ASD boys (Figure 2A). In female participants, there was a significant increase between baseline and follow-up in the cortical conduction velocity (T=2.809, p<0.05) but not a significant difference overall between ASD and the control group (T=-1.712, p=0.096, n.s.). In the white matter ROIs in male participants, there was neither a significant change over time (T=1.452, p=0.15, n.s.) nor a significant difference between ASD and the control group (T=0.225, p=0.82 n.s.), whereas in the female participants, there was a significant positive change between baseline and follow-up (T=3.502, p<0.01) but the amount of change did not differ between the ASD and the control group females(T=0.087, p=0.93, n.s.).

**Figure 2:**
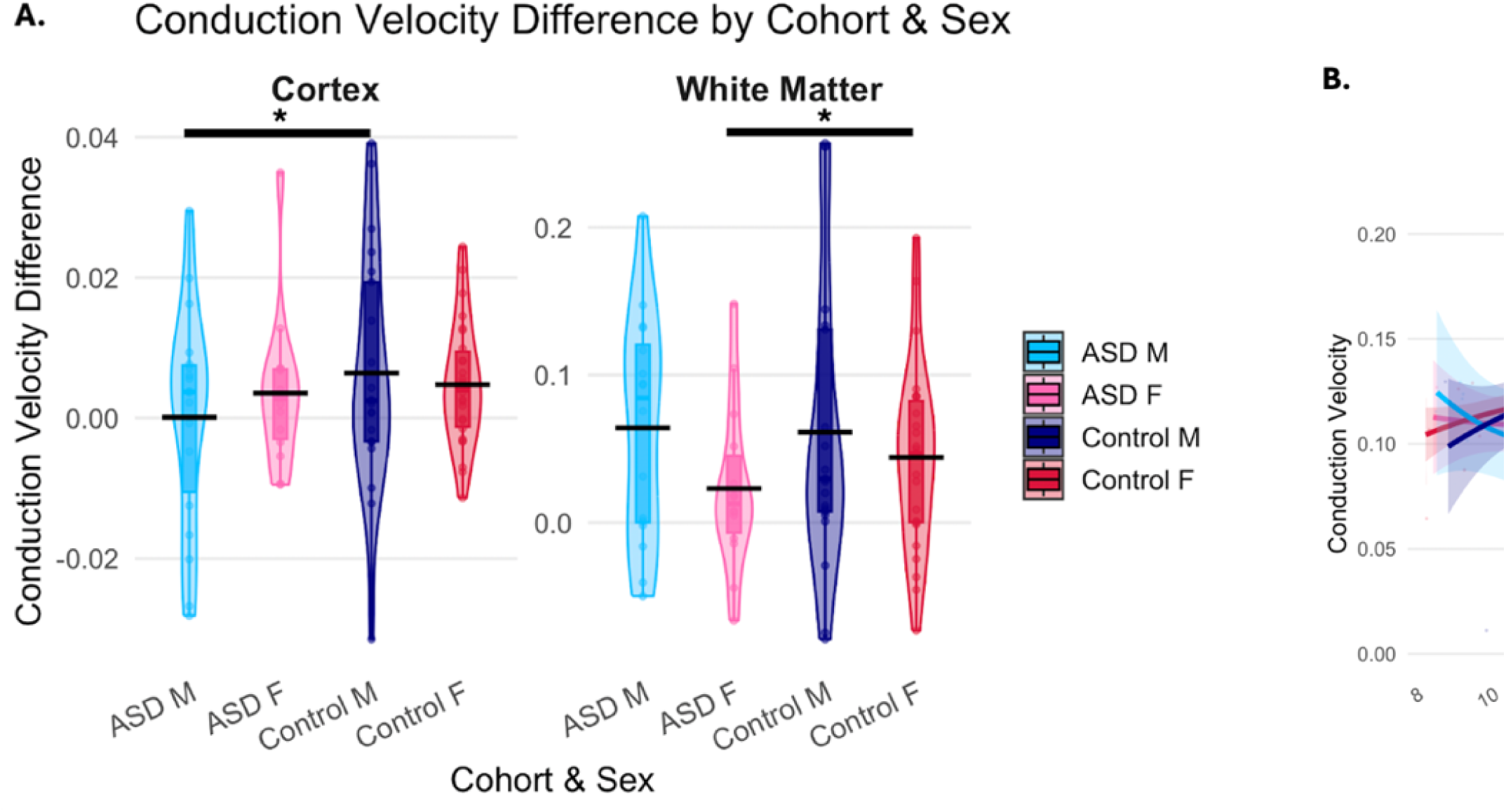
Violin plots showing mean change (Follow-up – Baseline) in ACV in cortex and WM (A). Plots showing ACV as a function of participant age in both cortex and WM (B).

**Figure 3:**
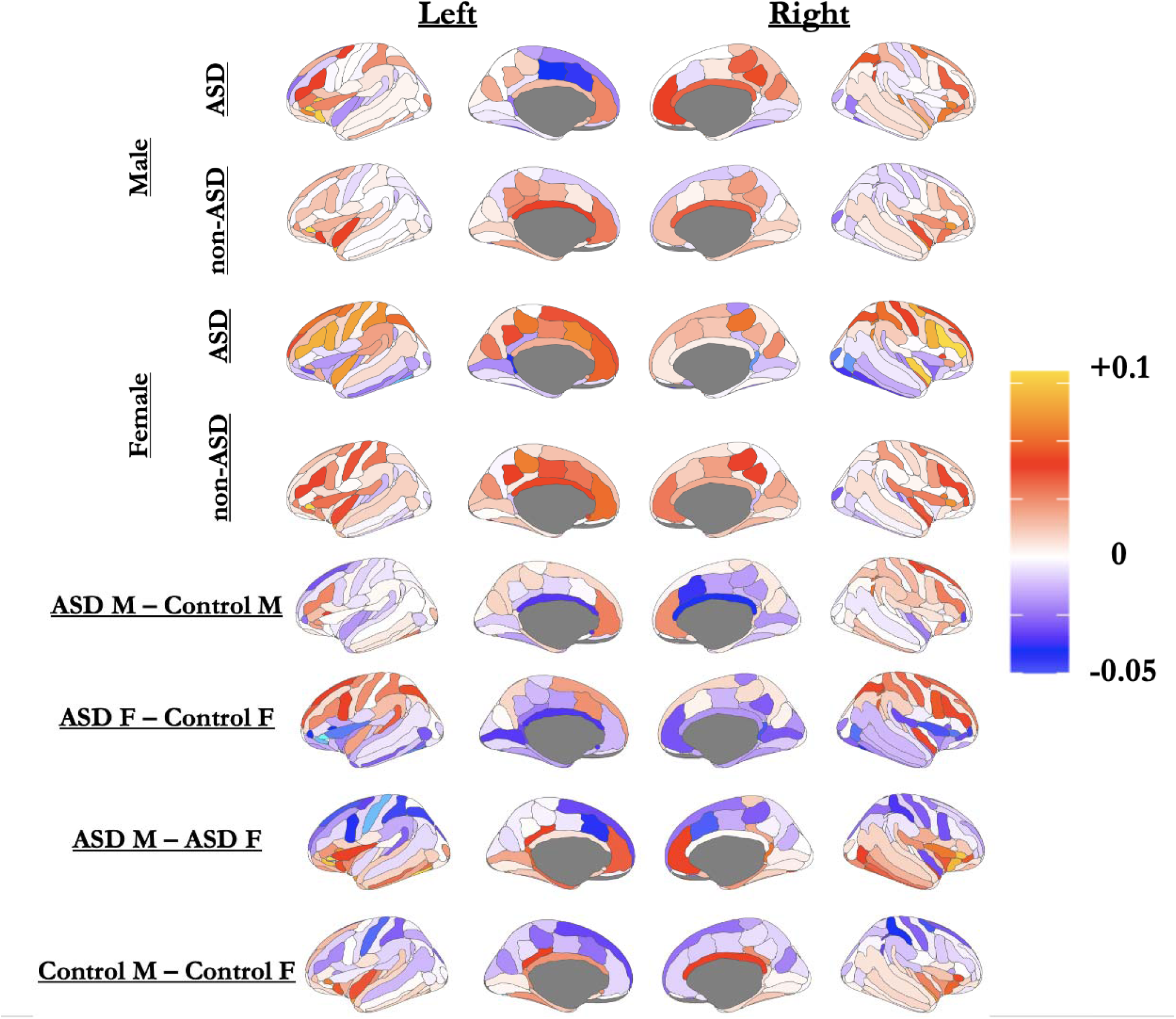
Mean change (Follow-up – Baseline) in each ROI in the 164 Destrieux Cortical Atlas, separated by diagnosis and sex. Red values indicate increases and blue values indicate decreases. Difference maps are displayed at bottom showing differences in rates of change (Follow-up – Baseline) among groups, with the latter labeled group subtracted from the former (i.e,. ASD M minus ASD F).

**Figure 4:**
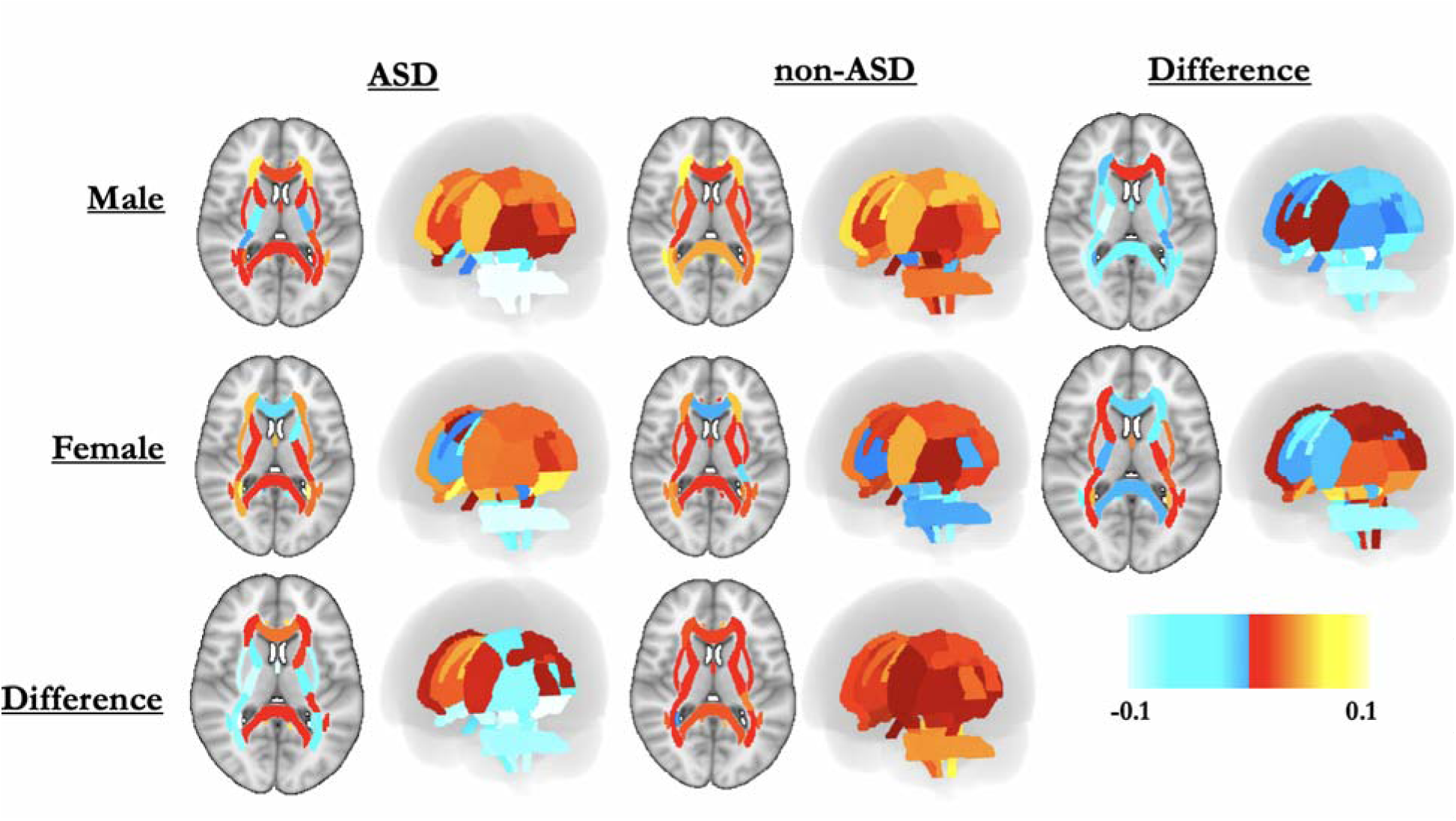
Mean change (Follow-up – Baseline) in each ROI in the 48 JHU WM Atlas, separated by diagnosis and sex. Red values indicate increases and blue values indicate decreases. Difference maps are displayed in the bottom row and right column showing differences in rates of change (Follow-up – Baseline) among groups, with the latter labeled group subtracted from the former (i.e., Male minus Female for columns, and ASD minus non-ASD for rows).

### Experiment 2

We additionally ran a mediation analysis to test whether pubertal development (PDS) explains the relationship between age and conduction velocity. The results showed a significant indirect effect of age on conduction velocity through PDS (ACME = 6.49e-05, p = 0.016), meaning that part of the effect of age is carried through pubertal status. The direct effect of age (not through PDS) was not significant (ADE = 5.24e-05, p = 0.200). Overall, the total effect of age on conduction velocity was significant (p < 0.001), and about 55% of that effect was mediated by PDS (p = 0.016).

Building on the observed longitudinal changes in neuroimaging metrics, we next analyzed behavioral and cognitive data within the ASD cohort to investigate how these functional measures evolved over time. To better understand potential sex-specific developmental trajectories, all analyses were stratified by sex. Behavioral score ranges by subscale, separated by timepoint and sex are visualized in Figure 5. We additionally examined longitudinal behavioral and cognitive changes across the full cohort, including both ASD and control participants; however, many metrics showed minimal change in the control group, likely due to floor effects where baseline scores were already within the normal range, leaving little room for measurable improvement. Behavioral data from the entire cohort of ASD and non-ASD participants is presented in Supplementary Figure 1.

**Figure 5:**
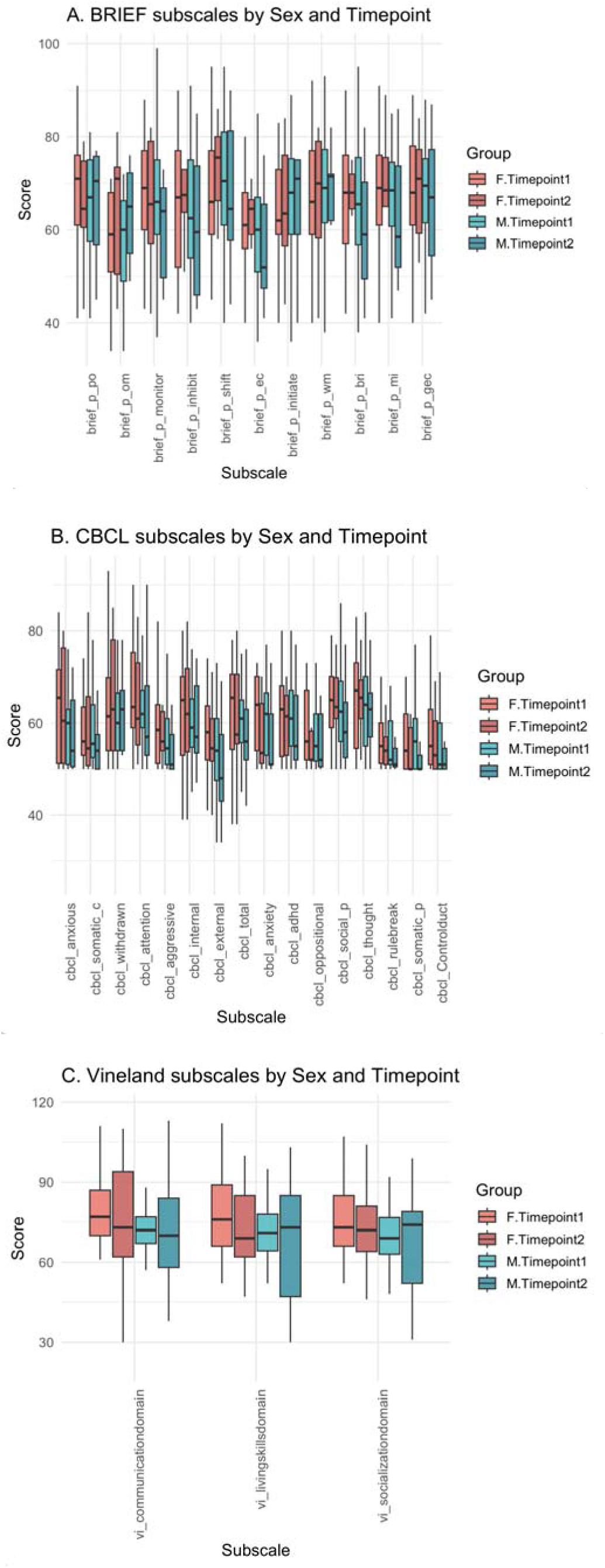
Longitudinal behavioral and cognitive subscale scores by sex in the ASD cohort. Boxplots display scores across multiple subscales at Timepoint 1 and Timepoint 2, separated by sex (female in red, male in blue) and test (BRIEF: A, CBCL: B, & Vineland-II: C). Each subscale represents a domain from behavioral (e.g., CBCL, BRIEF), cognitive (e.g., IQ composites), or adaptive functioning assessments.

### Experiment 3

To examine behavioral changes across time and potential sex differences in our ASD cohort, we fit linear mixed-effects models predicting each subscale score using sex, timepoint (first or second wave), and their interaction while adjusting for age, puberty development (using PDS scores), IQ, total brain volume, and data collection site, with a Benjamini and Hochberg FEW correction across all models. Significant sex x timepoint interactions emerged for several key behavioral domains, including CBCL total problems (p = 0.003), CBCL withdrawn (p = 0.010), CBCL Anxious/Depressed (p = 0.011), and CBCL internalizing (p = 0.016). These interactions suggest that changes in these subscales over time differed between males and females, with females exhibiting greater increases or sustained elevations in internalizing traits relative to males (Fig. 6). Although the main effect of timepoint was generally nonsignificant across most subscales, the effect of sex was significant for CBCL anxiety, with females demonstrating marginally higher scores than males. The covariates of age and pubertal status were significant across multiple domains. The full table of p-value results for all behavioral metrics is listed in Supplementary Table 2.

**Figure 6:**
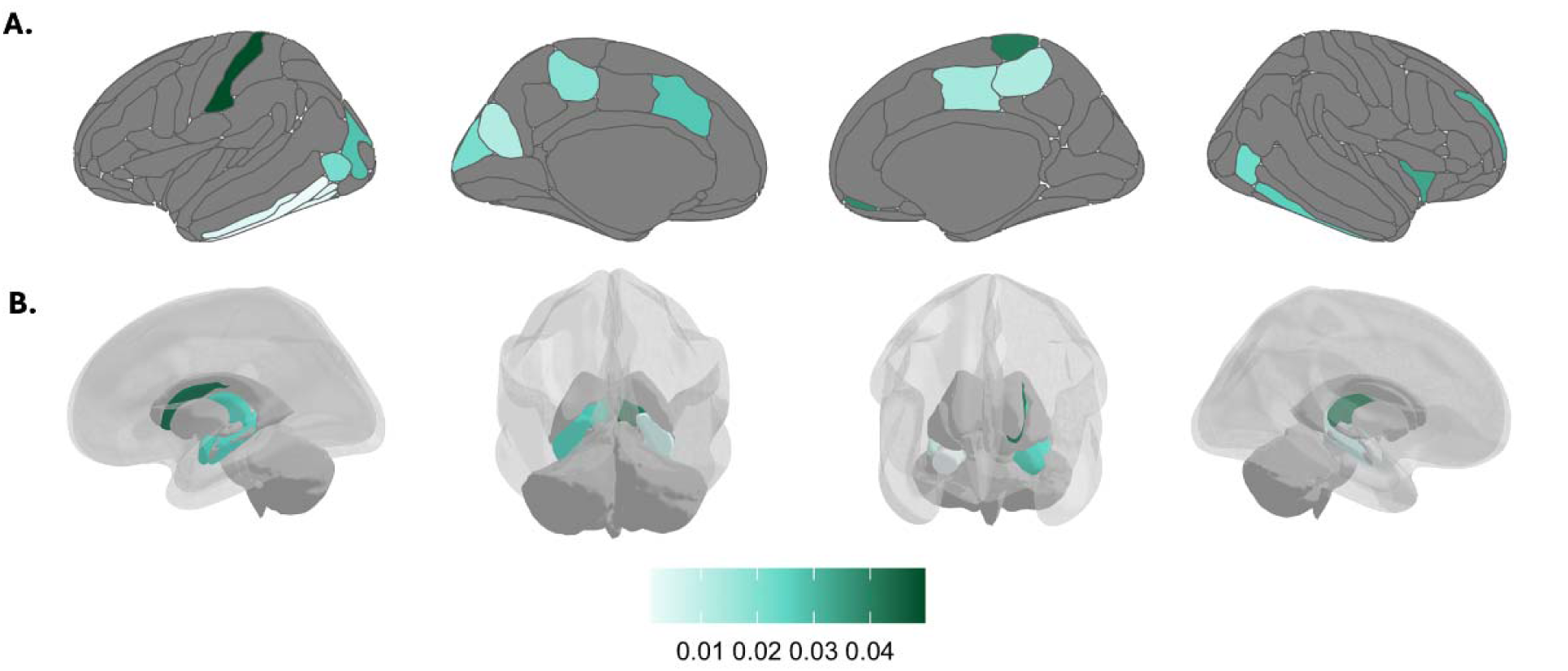
Surface renderings of significant associations between conduction velocity and CBCL Anxious/Depressed scores. A. Cortical regions shown on inflated surfaces. B. Subcortical regions visualized using 3D volumetric renderings within a transparent brain. Warmer colors represent stronger positive effect estimates, with the intensity of the shading corresponding to the magnitude of the association (see color bar). The model included age, sex, pubertal status (PDS), IQ, total brain volume, site, and timepoint as covariates.

To investigate the relationship between these behavioral and neuroimaging results, we ran linear mixed-effects models within each ROI to examine the association between cognitive variables and g-ratio and conduction velocity. Each model controlled for age, sex, IQ, ICV, site, and timepoint. Minimal significant associations were seen when looking at the complete cohort (controls and ASD participants), but several associations were seen when looking solely at the ASD cohort. Notably, there were 28 ROIs with significant associations between conduction velocity and CBCL Anxious/Depressed before Benjamini-Hochberg correction and 11 ROIs after correction.

Given the independent significant findings on the CBCL Anxious/Depressed subscale, both in the sex-by-timepoint interaction models and in its association with conduction velocity, we conducted a deeper analysis focused specifically on this measure. As shown in Figure 6, significant positive associations between conduction velocity and CBCL Anxious/Depressed scores were observed across several cortical regions. These included the left mid-anterior cingulate, cuneus, middle occipital, postcentral, and inferior temporal gyri. Darker colors on the cortical surface map reflect stronger effect estimates, with the left inferior temporal gyrus showing the most significant association (p = 0.0011). All models included age, sex, diagnosis, pubertal status, IQ, total brain volume, site, and timepoint as covariates.

Figure 7 illustrates brain regions with significant interaction effects between conduction velocity and CBCL Anxious/Depressed scores. The strongest effects were observed in the left postcentral gyrus (β = 0.0497), left caudate (0.0448), and right thalamus (0.0445), followed closely by the right paracentral gyrus/sulcus (0.0416) and right suborbital sulcus (0.0383). These associations spanned cortical, subcortical, and white matter regions, underscoring the distributed nature of neural correlates linked to anxiety symptoms.

**Figure 7:**
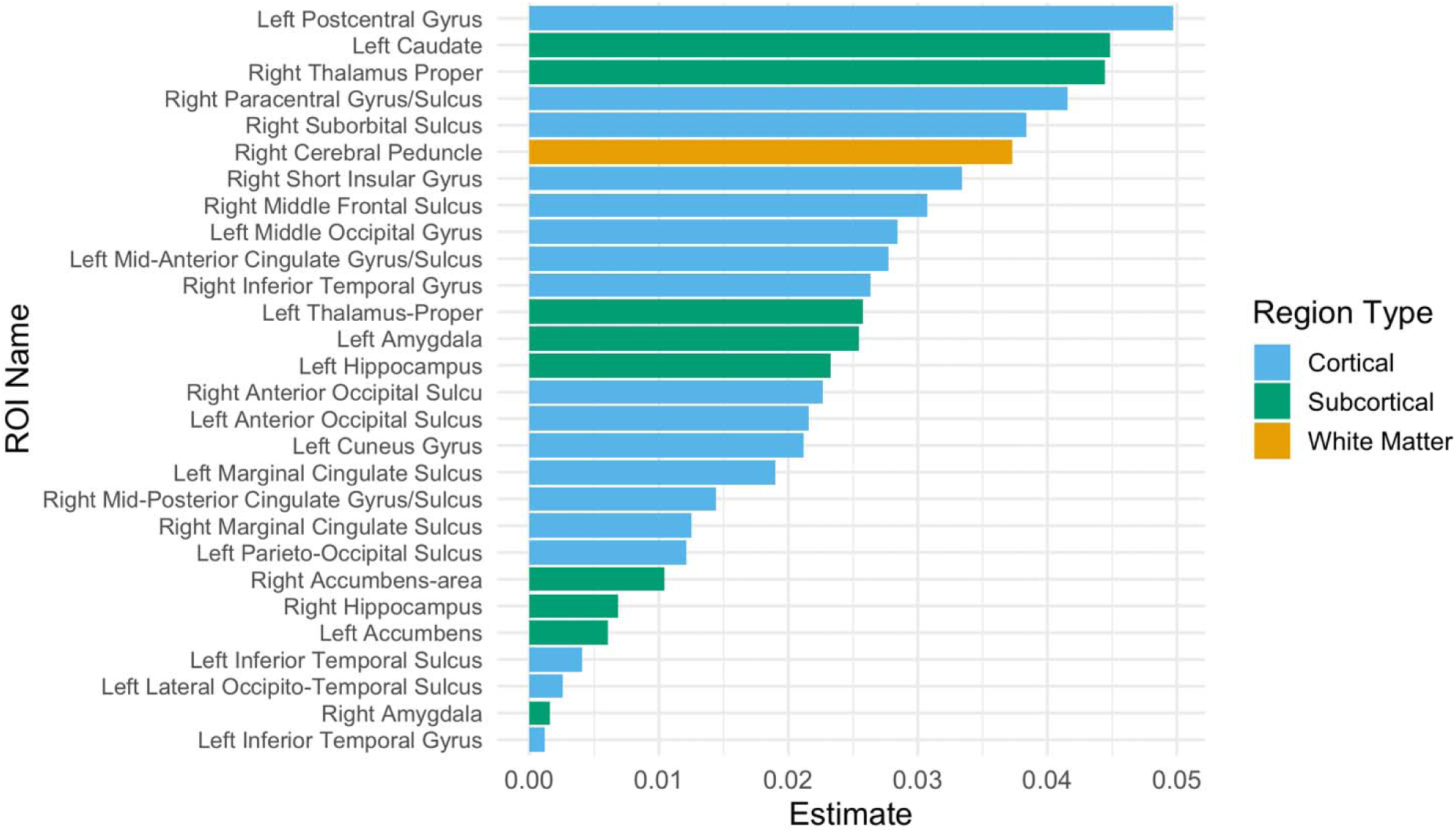
Brain regions showing significant interactions with CBCL Anxious/Depressed scores. Bar plot depicts regions of interest (ROIs) that showed significant interaction effects in a linear model examining CBCL Anxious/Depressed scores as a function of conduction velocity and regional brain structure. ROIs are color-coded by region type: cortical (blue), subcortical (green), and white matter (orange).

## Discussion

Our findings from longitudinal neuroimaging data are consistent with prior research demonstrating reduced cortical axonal conduction velocity in autistic individuals across development, while also showing that cortical and WM conduction velocity generally increases over time in both ASD and non-ASD participants. This suggests that, despite early differences, the overall trajectory of axonal maturation may follow a similar, though slower in ASD, developmental pattern across groups. However, the observation of reduced rates of cortical conduction velocity development in ASD males and a reduced rate of axonal conduction velocity development in ASD females points to sex-specific patterns of neurodevelopmental disruption. The strong mediating effect of pubertal development suggests sex-specific hormonal changes drive similar, but sex-specific, patterns of neuronal change. Girls showed faster rates of development in the frontal and parietal cortex, while boys, particularly those with ASD, showed much higher insular cortex development. Insular cortex development in boys may underlie the greatly improved behavioral and cognitive metrics experienced by the autistic boys in adolescence as the insular cortex is a well-established locus of social and emotional regulation(Gogolla, 2017; Lamm & Singer, 2010). The insula has previously been reported to be a major area associated with ASD behavioral manifestations (Caria & De Falco, 2015; Ebisch et al., 2011; Nomi et al., 2019). Such differences may contribute to the variability in trait presentation and cognitive outcomes observed in autistic males and females.

Findings from longitudinal behavioral analyses likewise emphasize the role of sex and the effect of puberty on development in the ASD cohort. While overall changes over time were modest, significant sex-by-timepoint interactions emerged for several internalizing subscales, most notably CBCL Anxious/Depressed. Females showed greater elevations in anxious and withdrawn behaviors over time as compared to males, supporting the existing literature suggesting that girls with autism are more likely to present with internalizing traits that often go underrecognized as they age (Mandy et al., 2012; Solomon et al., 2012). The CBCL Anxious/Depressed subscale was specially developed to evaluate internalizing traits and differs from the related subscale, CBCL Anxiety Problems, which captures more externally reported anxiety symptoms typically associated with treatment-seeking behaviors (Knepley et al., 2019).

Importantly, high scores on the CBCL Anxious/Depressed subscale were a marker of internalizing behavior and predicted scores on a range of other behavioral domains, even after adjusting for sex, timepoint, age, IQ, and site. This result suggests that anxiety could be a central organizing feature of behavioral variability in ASD, particularly in females. One possibility is that the manifestation of autism in girls is shaped in part by underlying anxiety that is not always expressed in typical ways, potentially contributing to misdiagnosis or late diagnosis.

Neuroimaging results further support this interpretation. CBCL Anxious/Depressed scores were associated with conduction velocity in more regions than any other behavioral metric, with significant associations concentrated in white matter regions previously shown to differ between ASD and non-ASD participants. Notably, these associations were strongest in females, suggesting a neurologically based interaction that may amplify anxiety symptoms during development. A key region, the Left Postcentral Gyrus, had both the strongest association with CBCL Anxious/Depressed scores (Fig. 6 & 7) and the greatest change in ASD girls (Fig. 3) that was additionally the greatest differential in slope between ASD boys and girls (Fig. 3). The sex-specific neurobiological underpinnings of anxiety may contribute to a cascading effect, influencing broader behavioral profiles and potentially masking or altering how autism presents in females.

Further research is needed to fully understand the neurobiological underpinnings of behavioral variability in ASD. In particular, longitudinal frameworks beginning in infancy have the potential to reveal earlier markers and clarify developmental trajectories seen in this study. Future work should continue to build toward biomarkers of ASD and reflect the broad range of variability within the condition, particularly its differences in girls versus boys. Moreover, these biomarkers should be used to enhance individualized support and early diagnostic abilities rather than pathologize the condition or enforce narrow definitions of typical development. In doing so, this work has the potential to inform more reliable diagnostic protocols and personalized interventions that account for the neurological basis of the condition.

## Data Availability

All de-linked phenotypic, neuroimaging, EEG, and genetics data from ACE GENDAAR has been shared with the NIMH Data Archive (NDA), beginning 11/23/2012, under study collection ID 2021 where they are available for download and re-analysis by NDA-approved investigators.

https://nda.nih.gov/

## Acknowledgements

This work was performed on behalf of the GENDAAR Consortium (NIH R01 MH100028), and we thank all of our collaborating colleagues, the study participants, and their family members. We would like to specifically acknowledge the contributions of Anna Kresse, MPH; Megha Santhosh, MHA; Désirée Lussier-Lévesque, PhD; Emily Neuhaus, PhD; Katy Ankenman, MSW; Jessica Benton, MA; and Rachel Fung, BS. The grant sponsors had no role in study design, data collection and analysis, decision to publish, or preparation of the manuscript.

## Supplementary Materials

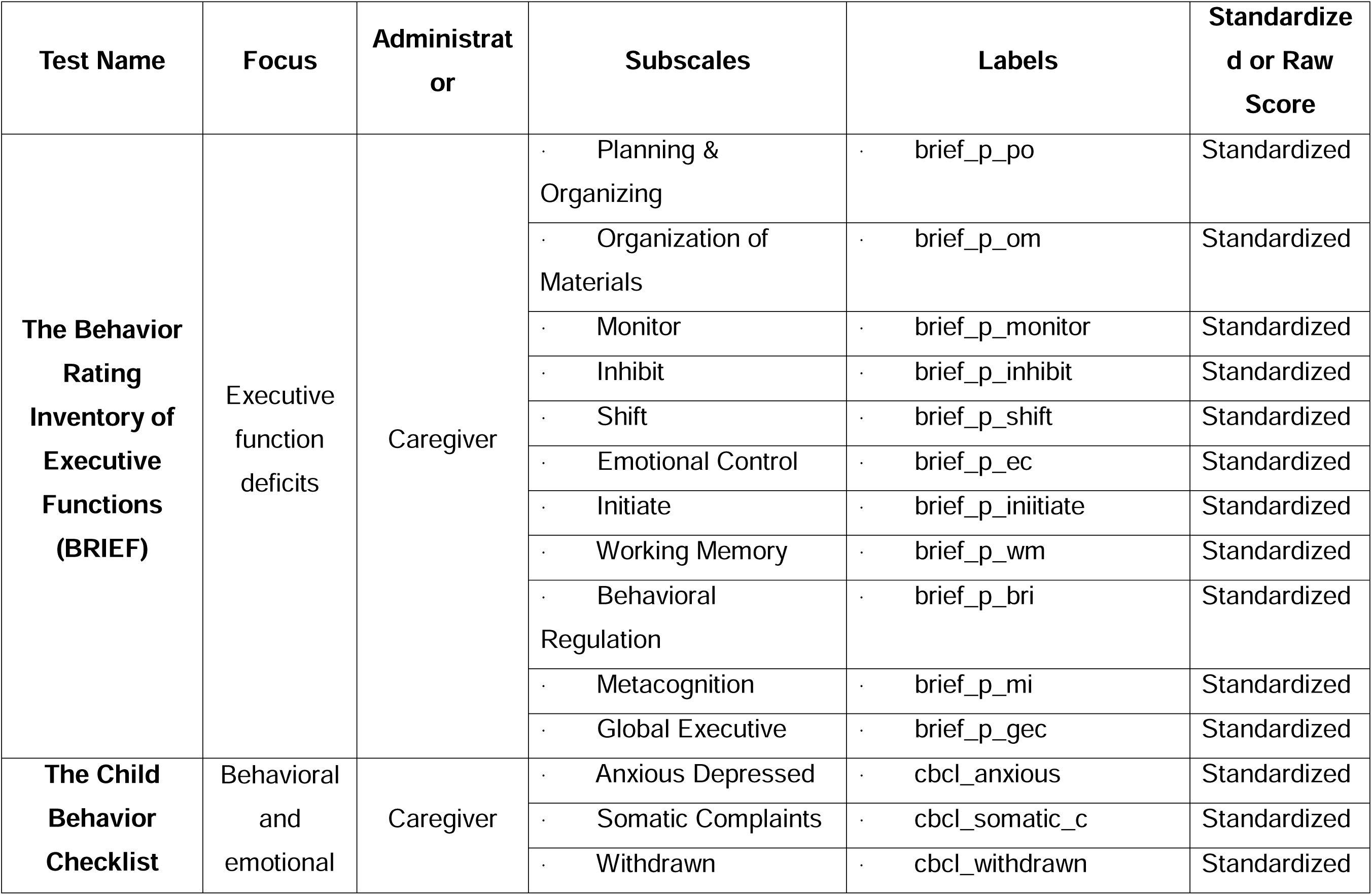
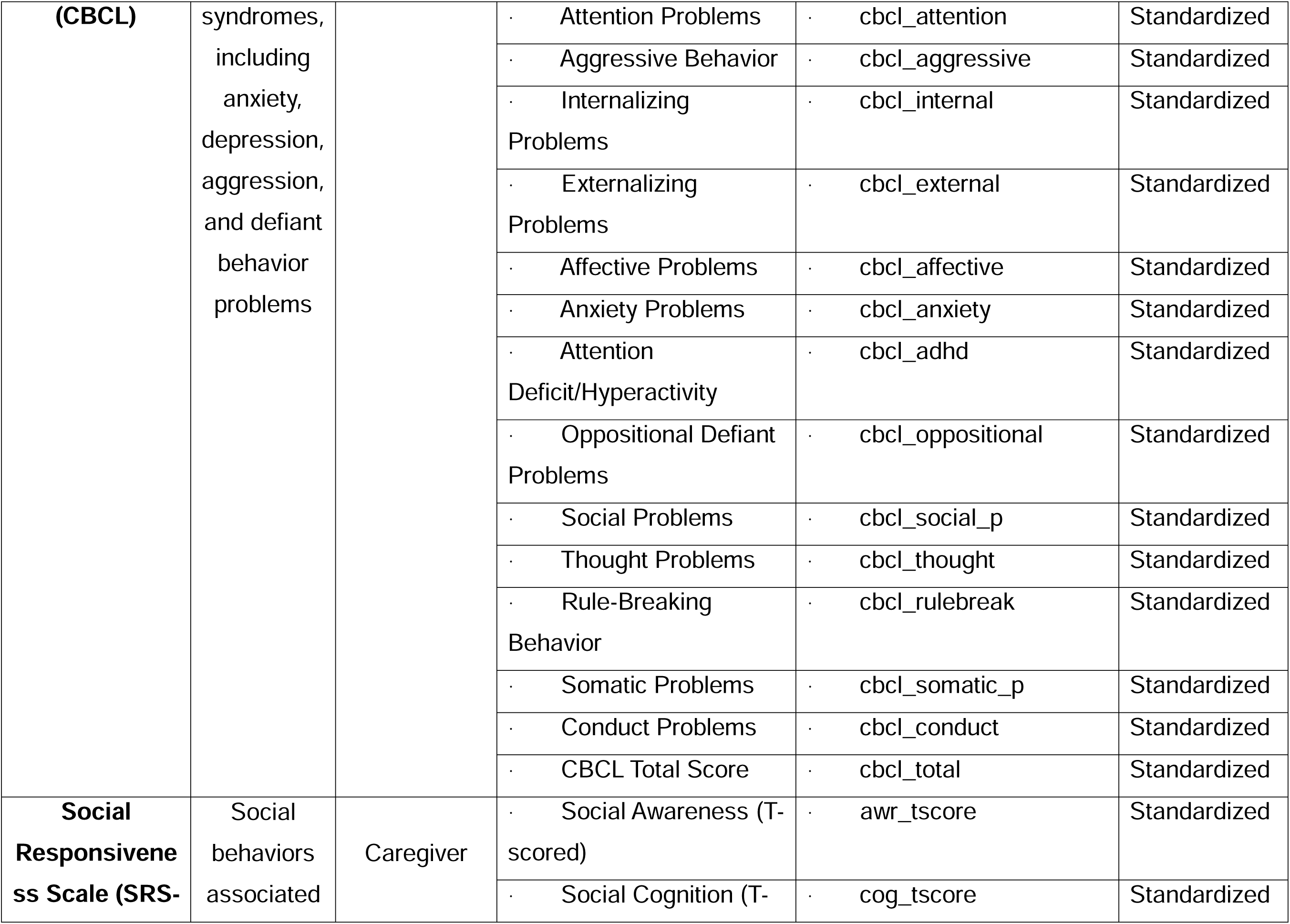
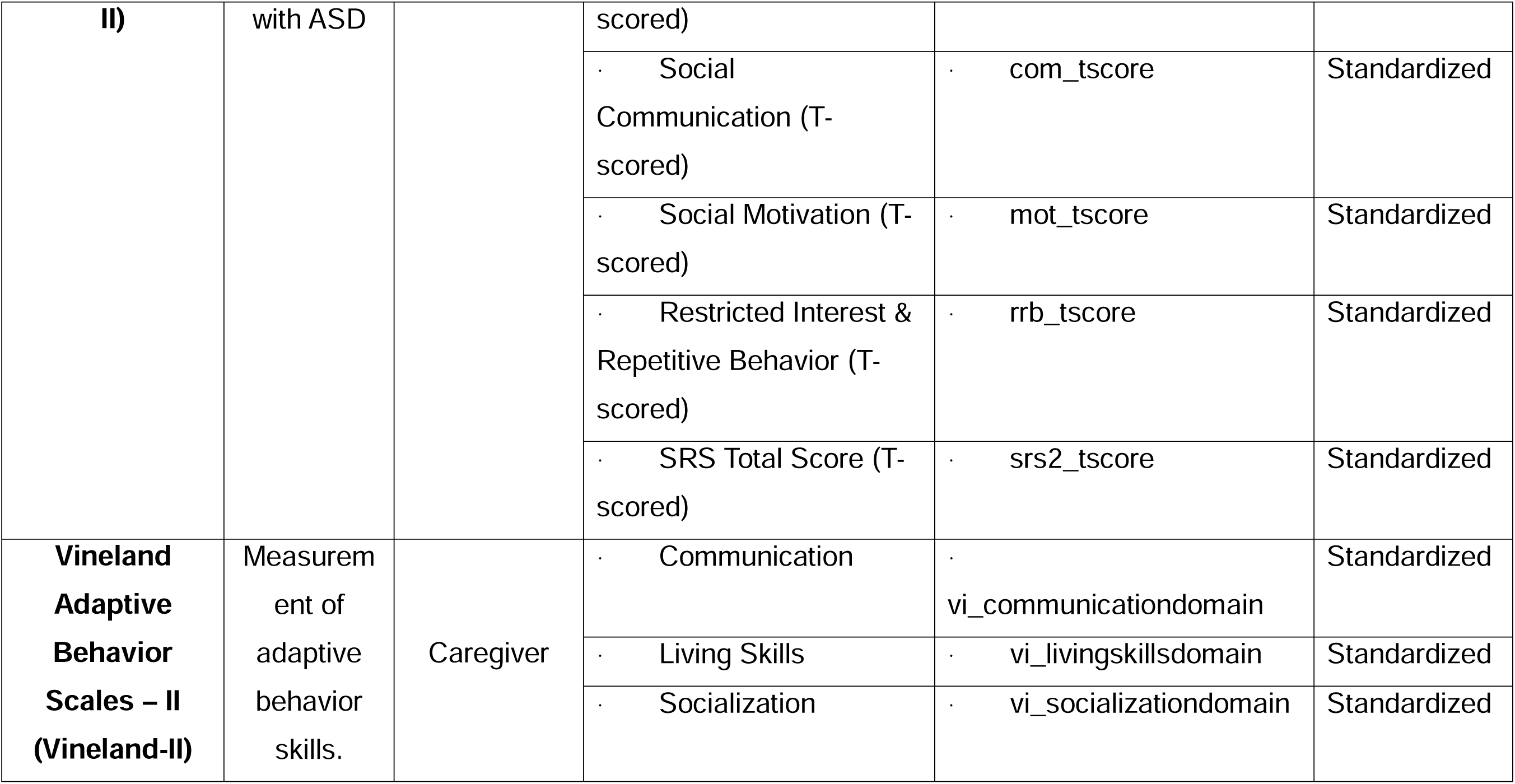

---

**Figure S1.**
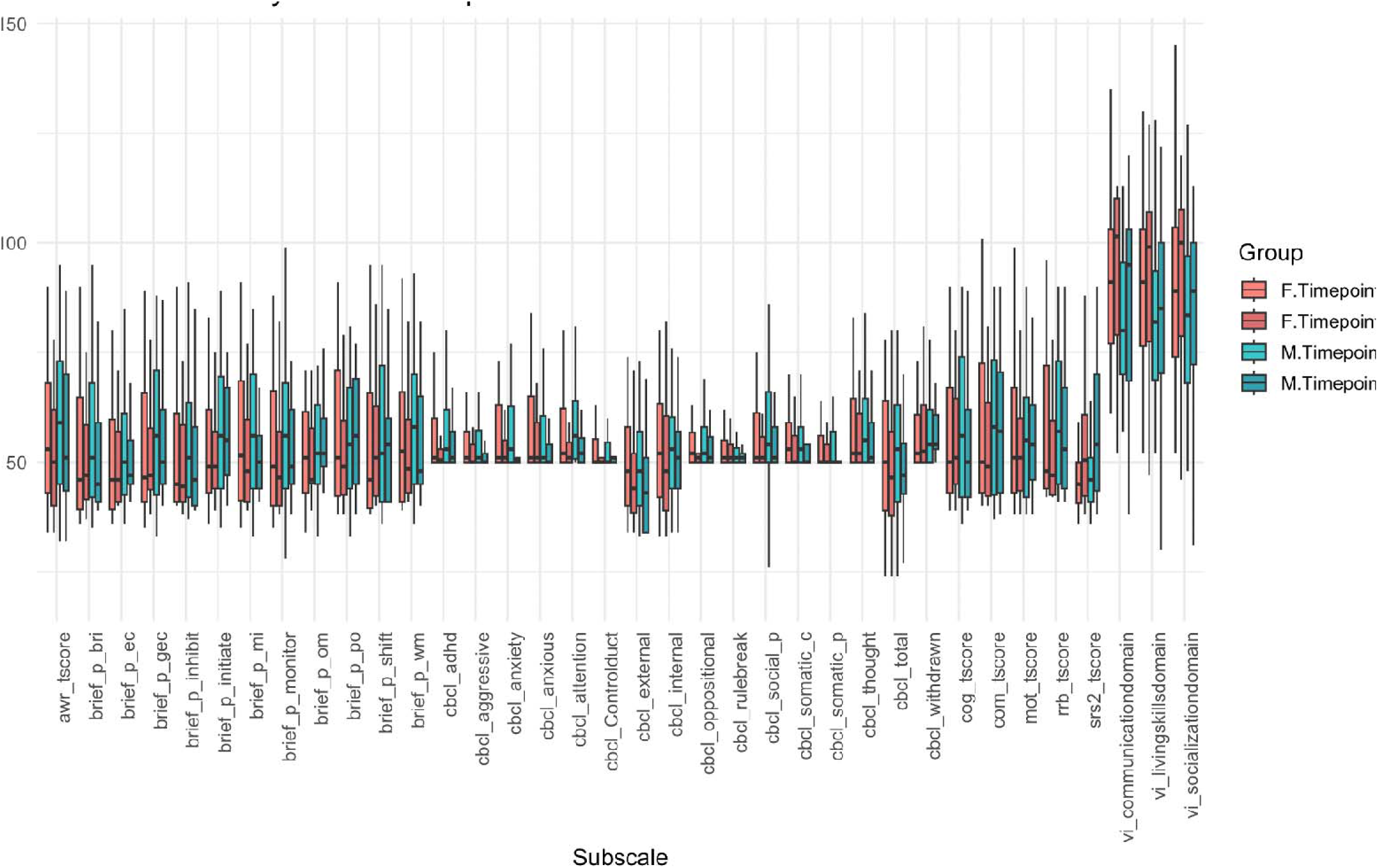
Behavioral and cognitive subscale scores by sex and timepoint across the full cohort. Boxplots show the distribution of scores on standardized behavioral and cognitive subscales at Timepoint 1 and Timepoint 2, stratified by sex (female in red, male in blue). The full cohort includes both autistic and neurotypical participants. Subscales span domains from instruments such as the BRIEF, CBCL, and Vineland. This visualization captures developmental trends and variability across groups over time.

