## Supplemental Table 1 for "Sex-specific Axonal Conduction Velocity Development Drives Differential Changes in Frontal, Parietal, and Insular Cortices in Autism Spectrum Disorder"

| **Test Name** | **Focus** | **Administrator** | **Subscales** | **Labels** | **Standardized or Raw Score** |
| --- | --- | --- | --- | --- | --- |
| **The Behavior Rating Inventory of Executive Functions (BRIEF)** | Executive function deficits | Caregiver | ·       Planning & Organizing | ·       brief_p_po | Standardized |
|  |  |  | ·       Organization of Materials | ·       brief_p_om | Standardized |
|  |  |  | ·       Monitor | ·       brief_p_monitor | Standardized |
|  |  |  | ·       Inhibit | ·       brief_p_inhibit | Standardized |
|  |  |  | ·       Shift | ·       brief_p_shift | Standardized |
|  |  |  | ·       Emotional Control | ·       brief_p_ec | Standardized |
|  |  |  | ·       Initiate | ·       brief_p_iniitiate | Standardized |
|  |  |  | ·       Working Memory | ·       brief_p_wm | Standardized |
|  |  |  | ·       Behavioral Regulation | ·       brief_p_bri | Standardized |
|  |  |  | ·       Metacognition | ·       brief_p_mi | Standardized |
|  |  |  | ·       Global Executive | ·       brief_p_gec | Standardized |
| **The Child Behavior Checklist (CBCL)** | Behavioral and emotional syndromes, including anxiety, depression, aggression, and defiant behavior problems | Caregiver | ·       Anxious Depressed | ·       cbcl_anxious | Standardized |
|  |  |  | ·       Somatic Complaints | ·       cbcl_somatic_c | Standardized |
|  |  |  | ·       Withdrawn | ·       cbcl_withdrawn | Standardized |
|  |  |  | ·       Attention Problems | ·       cbcl_attention | Standardized |
|  |  |  | ·       Aggressive Behavior | ·       cbcl_aggressive | Standardized |
|  |  |  | ·       Internalizing Problems | ·       cbcl_internal | Standardized |
|  |  |  | ·       Externalizing Problems | ·       cbcl_external | Standardized |
|  |  |  | ·       Affective Problems | ·       cbcl_affective | Standardized |
|  |  |  | ·       Anxiety Problems | ·       cbcl_anxiety | Standardized |
|  |  |  | ·       Attention Deficit/Hyperactivity | ·       cbcl_adhd | Standardized |
|  |  |  | ·       Oppositional Defiant Problems | ·       cbcl_oppositional | Standardized |
|  |  |  | ·       Social Problems | ·       cbcl_social_p | Standardized |
|  |  |  | ·       Thought Problems | ·       cbcl_thought | Standardized |
|  |  |  | ·       Rule-Breaking Behavior | ·       cbcl_rulebreak | Standardized |
|  |  |  | ·       Somatic Problems | ·       cbcl_somatic_p | Standardized |
|  |  |  | ·       Conduct Problems | ·       cbcl_conduct | Standardized |
|  |  |  | ·       CBCL Total Score | ·       cbcl_total | Standardized |
| **Social Responsiveness Scale (SRS-II)** | Social behaviors associated with ASD | Caregiver | ·       Social Awareness (T-scored) | ·       awr_tscore | Standardized |
|  |  |  | ·       Social Cognition (T-scored) | ·       cog_tscore | Standardized |
|  |  |  | ·       Social Communication (T-scored) | ·       com_tscore | Standardized |
|  |  |  | ·       Social Motivation (T-scored) | ·       mot_tscore | Standardized |
|  |  |  | ·       Restricted Interest & Repetitive Behavior (T-scored) | ·       rrb_tscore | Standardized |
|  |  |  | ·       SRS Total Score (T-scored) | ·       srs2_tscore | Standardized |
| **Vineland Adaptive Behavior Scales – II (Vineland-II)** | Measurement of adaptive behavior skills. | Caregiver | ·       Communication | ·       vi_communicationdomain | Standardized |
|  |  |  | ·       Living Skills | ·       vi_livingskillsdomain | Standardized |
|  |  |  | ·       Socialization | ·       vi_socializationdomain | Standardized |
